# The epidemiological characteristics of the primary health care based COVID-19 swabbed persons in Qatar, March- October 2020

**DOI:** 10.1101/2021.02.17.21251722

**Authors:** Mohamed Ghaith Al-Kuwari, Mariam Ali Abdulmalik, Ahmad Haj Bakri, Mujeeb Chettiyam Kandy, Maha Youssef Abdulla, John Gibb

**Affiliations:** Primary Health Care Corporation, Qatar

**Keywords:** COVID-19, Qatar, Primary care, Epidemiology

## Abstract

**Background:** In March 2020, Qatar started reporting increased numbers of COVID-19 positive cases. National preventive measures were put in place and testing plan has been developed to respond to the pandemic with the primary health care as the main provider. The study aims to describe the epidemiological characteristics of COVID-19 at the primary health care level in Qatar and to examine the factors associated with the positivity rate.

**Method:** Retrospective data analysis for all the cases screened for COVID-19 at the primary health care level in Qatar between the 11th of March and 31st of October 2020. The study analyzed the demographic characteristics of the tested persons and non-communicable disease burden, positivity rate by month, nationality, and age-group, factors associated with the positivity rate.

**Results:** Between the 11th of March and the 31st of October 2020, 285,352 persons were tested for SARS-CoV-2, with a median age (IQR) of 32 (22-43) years. 59.9% were from the Middle East and North Africa region and 29.6% originally from Asia. Overall, among them, 11.2% had diabetes mellitus and 11.4% had hypertension. The epidemiological curve showed a steep increase in positivity rate from March till June 2020 at the highest rate of 21.4% in June 2020. The highest positivity rate was observed among Asian males at 20.3%. The positivity rates were almost the same among the tested persons for SARS-CoV-2 in the main three age-groups (0-18, 19-39, 40-59) at 13%, 14.5%, and 14.0%, respectively. In a multi regression model, being a male was associated with a higher risk (OR 1.16; 95% CI 1.13 to 1.18). Persons originally from Asia were at higher risk than those originally from the Middle East and North Africa (OR 1.5; 95% CI 147 to 1.54). COVID-19 infection was higher among those presenting with clinical symptoms than those asymptomatic (OR. 4.16; 95% CI 4.05 to 4.28).

**Conclusion:** The epidemic predominantly affected younger ages and males namely coming from Asia. At the primary health care level, COVID-19 infection rate was higher among those who presented with clinical symptoms. The scale-up of the testing at the primary health care level helped in detecting more cases and was reflected in a steady increase in the positivity rate to be flattened afterward.

## Introduction

In December 2019, a cluster of patients with unknown causes of pneumonia was reported in Wuhan, China.^1 2^ The samples’ viral genetic sequencing indicated a novel coronavirus. ^1^ The novel virus was named 2019 novel coronavirus (COVID-19) and 75-80% resemblance to SARS-CoV was confirmed. ^3^ It was presumed initially to be transmitted from animals to humans however, the virus has since spread rapidly through human to human transmission to the globe.^4–6^ As of December 29, 2020, approximately 79 million reported cases and over 1.7 million deaths have been reported globally. ^7^

The published epidemiological studies across several populations demonstrate substantial differences in the severity of infection and rates and in case fatality rates. ^8^ Hence, analyzing the transmission patterns among the population with unique demographics can improve the knowledge of the disease dynamic. Qatar is located in the Middle East peninsula with borders with the Arabian Gulf and Saudi Arabia. Qatar’s estimated population in July 2020 was almost 2.4 million with a unique population demographic profile in which the expatriate workforce constitutes almost 88% of the population. ^9^ The expatriate workforce nature in Qatar influences the male to female ratio with almost 75% male to 25% female, the population pyramid is condensed in the age groups 25-40 year, namely among males. ^10^ Evidence from other countries demonstrated that COVID-19 affects disproportionally males with poorer outcome among the older age group.^11 12^

Increased population movement and mobility made it possible for COVID-19 to spread more easily and faster. ^13^ In Qatar and a general restriction on all incoming international flights was implemented on the 31st of March 2020. The latter helped in halting almost all the visitors or residents into the country. In March 2020, Qatar started reporting increased numbers of COVID-19 positive cases. At that stage, national restrictions were put in place. The Ministry of Public Health in Qatar developed an emergency action plan to respond to the outbreak of COVID-19 with the Primary Health Care Corporation (PHCC) as the main component of that response.

Primary Health Care Corporation (PHCC), the main primary care provider in Qatar is serving 1.4 million individuals throughout a network of 27 primary health care centers covering all three main regions in the country (central, northern and western). PHCC responded rapidly to the pandemic by opening the first COVID-19 center for testing and holding on the 12th of March 2020. By end of March 2020, PHCC scaled up its testing capacity at the community level by designating 3 additional health centers for testing and holding and allocating at least one testing room in the remaining 23 health centers. The main objective of this study is to define the epidemiological characteristics of COVID-19 at the primary health care level in Qatar and to examine the factors associated with infectious rate. Understanding the epidemiological characteristics associated with COVID-19 infection at the primary health care level in Qatar will be beneficial in understanding the epidemiology in countries with unique demographic characteristics.

## Methods

Primary health care corporation by 11th of March 2020 had 4 designated health centers for COVID-19 testing in addition to at least one allocated room in each of the remaining 23 health centers for COVID-19 testing. All COVID-19 testing in Qatar was performed by Hamad medical corporation central lab, which is the public main secondary/ tertiary care provider in the state of Qatar and provides more than 85% of the inpatient bed capacity in the country.^14^ The test results are shared with the PHCC through the joint electronic medical record system between the two entities on an hourly basis. The Ministry of Public Health designed and implemented an effective contract tracing system. For each person who tested positive a thorough contact tracing was conducted by trained staff at the ministry of Public Health. ^14^

Between the 11th of March and the 31st of October 2020, the persons who were tested at PHCC testing sites were mainly persons presenting with influenza-like symptoms suggesting COVID-19 illness in addition to persons who have been in contact with COVID-19 patients. Nasopharyngeal and throat swabs were collected from the latter. To detect SARS-CoV-2 infection real-time RNA-PCR was utilized using TaqPath COVID-19 Combo Kit (Thermo Fisher Scientific, Waltham, Massachusetts, USA) or Cobas SARS-CoV-2 Test (Roche Diagnostics, Rotkreuz, Switzerland). These tests are proven to be highly sensitive and specific.^15 16^

The nationality of each tested person was established based on the official identification state card, the card is issued for all the residents and nationals in Qatar. Nationalities were grouped into four main groups based on the demographic distribution of ethnicity in Qatar for analysis purposes (Middle Eastern and North African, Asian, African, and others for Europeans, Americans and Australians, and New Zealanders). Demographic characteristics and burden of non-communicable diseases among the tested persons were derived from their electronic medical records. The age of the tested persons was categorized into four main groups (0-18, 19-39, 40-59, 60+).

A timeline of the positivity rate throughout the study period was created to study the trend of the epidemic at the primary health care level. Positivity rate was calculated among the different grouped nationalities and age-groups and persons presenting with symptoms. Multivariable logistic regression was used to determine the effect of age, symptoms presentation, and grouped nationality on capturing COVID-19 infection.

This study was conducted after securing all the necessary approvals from the PHCC scientific committee in response to the national and global public health emergency. There was no direct patient or public involvement, yet the key elements of the data were shared daily with the public.

## Results

Between the 11th of March and the 31st of October 2020, 285,352 persons were tested for SARS-CoV-2, with a median age (IQR) of 32 (22-43) years. Around 48.6% of those tested persons were in the age-group of 19-34 years followed by 24.3% and 20.3% of them falling into the age-group of 40-59 years and 0-18 years, respectively as shown in table No.1.

**Table No. 1:** Characteristics of the swabbed persons at the PHCC health centers between 11^th^ of March and 31^st^ Oct 2020

|  | Male n (%) | Female n (%) | Total n (%) |
| --- | --- | --- | --- |
| **Demography** |  |  |  |
| Mean age, years (SD) | 32.8 (17.1) | 31.9 (16.4) | 32.4 (16.8) |
| Median age, years (IQR) | 32 (21-44) | 32 (22- 42) | 32 (22- 43) |
| 0-18 years | 30,225 (20.3) | 27,694 (20.3) | 57,919 (20.3) |
| 19-39 years | 70,189 (47.2) | 69,208 (50.6) | 139,397 (48.6) |
| 40-59 years | 37,418 (25.2) | 32,026 (23.4) | 69,444 (24.3) |
| 60+ years | 10,798 (7.3) | 77,94 (5.7) | 19,592 (8.42) |
| **Nationality grouped** |  |  |  |
| MENA | 91,343 (61.5) | 79,552 (58.2) | 170,895 (59.9) |
| Asian | 43,095 (29.0) | 41,444 (30.31) | 84,539 (29.6) |
| African | 3,060 (2.1) | 4,969 (3.6) | 8,029 (2.8) |
| Others | 11,132 (7.5) | 10,757 (7.9) | 21,889 (7.8) |
| **Reason for swabbing** |  |  |  |
| Clinical suspicion | 40,482 (27.2) | 41,497 (30.6) | 81,979 (28.7) |
| Individual request | 16,689 (11.2) | 14,728 (10.8) | 31,417 (11.0) |
| Survey | 18,493 (12.4) | 16,049 (11.7) | 34,542 (12.1) |
| Contact tracing | 14,061 (9.5) | 15,715 (11.5) | 29,776 (10.4) |
| Port of entry | 15,364 (10.3) | 14,442 (10.7) | 29,806 (10.5) |
| Healthcare routine check up | 2,616 (1.8) | 3,200 (2.3) | 5,816 (2.0) |
| Others- Not specified | 11,339 (7.6) | 10,488 (7.7) | 21,827 (7.6) |
| Missing | 20,603 (15.0) | 29,586 (19.9) | 50,189 (17.6) |
| **Symptoms presentation** |  |  |  |
| Symptomatic | 33,908 (28.7) | 35,631 (30.9) | 69,539 (29.7) |
| Asymptomatic | 84,416 (71.3) | 79,775 (69.1) | 164,191 (70.3) |
| **Comorbidities** |  |  |  |
| Diabetes mellitus | 17,788 (12.0) | 14,031 (10.3) | 31,819 (11.2) |
| Cardiovascular disease | 4,865 (3.3) | 1,857 (1.4) | 6,722 (2.5) |
| Hypertension | 18,568 (12.5) | 13,880 (10.1) | 32,448 (11.4) |
| Asthma/COPD | 12,754 (8.6) | 11,771 (8.6) | 24,525 (8.6) |
| Dyslipidemia | 16,254 (10.9) | 13,146 (9.6) | 29,400 (10.3) |

Persons originally from the Middle East and Northern Africa region constituted more than half of the test persons for SARS-CoV-2 with 59.9% followed by persons originally from Asia with 29.6% as indicated in table No.1.

The main reason for swabbing was clinical suspicion with 28.7% of persons being swabbed due to influenza-like symptoms and severe acute respiratory infection. Random swabbing surveys among the clusters of high-risk population and contact tracing swabbing for the persons who have been identified as in contact with positive COVID-19 cases based on the criteria published by the US Centers for Disease Control and Prevention were among the main reasons for swabbing with 12.1% and 10.4%, respectively as shown in table No.1.

Among the persons who were tested for SARS-CoV-2 at the PHCC testing sites, the most common comorbidities were diabetes mellitus (11.2%), hypertension (11.4%), Asthma and COPD (8.6%), and cardiovascular diseases (2.5%). Diabetes mellitus was slightly higher among males than that among females (12.0% vs. 10.1).

At the beginning of March 2020 when the COVID-19 cases started rising in Qatar, the PHCC designated four health centers as exclusive COVID-19 testing and holding centers. Also, one testing room at least was allocated within all other remaining health centers. The positivity rate was calculated per month as demonstrated in graph No.2. The epidemiological curve showed a steep increase in positivity rate as of March till June 2020, then there was an increase again in the positivity rate in June and August aligned with entering the phase one and two of lifting restrictions. The positivity rate started to decrease as of October 2020.

The grouped nationalities with the highest positivity rates were Asian (17.9%), Middle Eastern and North African (12.6%), and African (12.6%) as shown in table No.2. Asian males had the highest positivity rate at 20.3%. The positivity rates were almost the same among the tested persons for SARS-CoV-2 in the main three age-groups (0-18, 19-39, 40-59) at 13%, 14.5%, and 14.0%, respectively. Health centers located in the western region of Qatar had the highest positivity rate in comparison to remaining health centers located in the other two regions at 15.2%. Positivity rate was the highest among the persons who presented with clinical symptoms of influenza-like illness and severe acute respiratory infection at 20% as illustrated in table No.2.

**Table No. 2:** Positivity rate by nationality grouped, age-group, region and symptoms presentation

|  | Male n (%) | Female n (%) | Total n (%) |
| --- | --- | --- | --- |
| **Nationality-grouped** |  |  |  |
| MENA | 11,754 (12.9) | 9,830 (12.4) | 21,584 (12.6) |
| Asian | 87,37 (20.3) | 6,377 (15.4) | 15,114 (17.9) |
| African | 412 (13.5) | 598 (12.3) | 1,010 (12.6) |
| Others | 785 (7.1) | 771 (7.7) | 1,556 (7.1) |
| **Age-group** |  |  |  |
| 0-18 | 3,789 (12.5) | 3,711 (13.4) | 7,500 (13.0) |
| 19-39 | 11,086 (15.8) | 9,144 (13.1) | 20,230 (14.5) |
| 40-59 | 5,730 (15.3) | 4,009 (12.5) | 9,739 (14.0) |
| 60+ | 1,083 (10.0) | 712 (9.1) | 1,795 (9.7) |
| **Region** |  |  |  |
| Central | 6,962 (14.7) | 5,817 (12.4) | 12,779 (13.5) |
| Northern | 7,935 (13.2) | 6,627 (12.6) | 14,562 (12.9) |
| Western | 6,791 (16.4) | 5,132 (13.9) | 11,923 (15.2) |
| **Symptoms presentation** |  |  |  |
| Asymptomatic | 4,982 (5.9) | 4,739 (5.9) | 9,721 (5.2) |
| Symptomatic | 7,352 (21.70) | 7,093 (19.9) | 14,445 (20.8) |

In a multivariable logistic regression model sex, nationality group, age-groups, and symptoms presentation were examined as exposure factors on capturing COVID-19 infection. Sex was found to have an effect with being a male was associated with a higher risk (OR 1.16; 95% CI 1.13 to 1.18). Nationality group had an effect on capturing COVID-19 infection with persons originally from Asia being at higher risk than those originally from the Middle East and North Africa (OR 1.5; 95% CI 147 to 1.54).

COVID-19 infection was higher among those presenting with clinical symptoms than those asymptomatic (OR. 4.16; 95% CI 4.05 to 4.28) as shown in table No.3. When the analysis was repeated for each exposure factor adjusting for the remaining almost same ORs were provided.

**Table No. 3:** Factors associated with the positivity rate (multi regression model)

| Variable | OR | 95% CI | P value |
| --- | --- | --- | --- |
| **Sex** |  |  |  |
| Female (reference) |  |  |  |
| Male | 1.16 | 1.13 to 1.18 | 0.000 |
| **Age-group** |  |  |  |
| 0 - 18 (reference) |  |  |  |
| 19-39 | 1.14 | 1.1 to 1.74 | 0.000 |
| 40-59 | 1.1 | 1.07 to 1.13 | 0.000 |
| 60+ | 0.72 | 0.68 to 0.76 | 0.000 |
| **Nationality-grouped** |  |  |  |
| MENA (reference) |  |  |  |
| Asian | 1.51 | 1.47 to 1.54 | 0.000 |
| African | 0.99 | 0.93 to 1.06 | 0.000 |
| Others | 0.53 | 0.50 to 0.55 | 0.000 |
| **Symptoms presentation** |  |  |  |
| Asymptomatic (reference) |  |  |  |
| Symptomatic | 4.16 | 4.05 to 4.28 | 0.000 |

## Discussion

This study provides the epidemiological characteristics of the SARS-CoV-2 outbreak at the primary health care level in Qatar where primary health care corporation operates, which adds a better understanding of the behavior of the pandemic in a unique demographic setting.

In Qatar, testing for SARC-CoV-2 started on the 5th of February 2020 with the first case identified on the 28th of February 2020, among returning travelers. During this time the infection had spread in many countries over the globe. Meanwhile, the State of Qatar established a national plan to respond to the anticipated COVID-19 outbreak, and testing for SARC-CoV-2 was scaled up as of March 2020 with the first cluster of cases of 300 was identified on the 8th of March 2020 among expatriate workers. The effective and restrictive case identification and contact tracing measures were probably the reasons behind the low number of cases up until the end of March 2020. During that time, a substantial number of returning travelers and nationals were identified to have COVID-19 infection.

Qatar took a series of public health measures in response to the spread of COVID-19. The measures included limiting incoming passenger flights into Doha through Hamad International Airport and allocating quarantine facilities for returning travels. Other measures were put in place gradually that promoted physical distancing, including closing retail stores and malls, suspending larger sports activities, and enforcing working from home for 80% of the public and private sectors. ^14^ The latter measures started to ease based on a phased approach for lifting restrictions as of the 15th of June 2020 onwards.

As of April, the number of daily cases and subsequently the positivity rate started to increase steadily at the community level, in part attributed to the scaled-up capacity of the PHCC in conducting screening tests and also to the expansion of the epidemic in the wider population. The positivity rate reached its highest level in June at 21.4%. The latter might be attributed to relaxing the restriction measures in Qatar by entering phase one of lifting restrictions on the 15th of June 2020 and due to the Eid Break, where people as part of the social behavior tend to gather for Eid celebration.

The epidemiological characteristics of the tested cases at the community level through PHCC testing sites showed that the majority of the tested cases were for persons coming from the Middle East and Northern Africa region at 59.9% followed by persons originally coming from Asia at 29.6%. The latter is a reflection of the distribution of the registered population at the primary health care services in Qatar, which is a reflection of the diversity and dynamics of the demographic distribution of the population in the state. In Qatar, 88% of the population are expatriates due to the workforce needs. ^2^ Persons who reported clinical symptoms of influenza-like-illness when conducting the COVID-19 test at the PHCC testing sites had a higher COVID-19 infection rate than those who were asymptomatic (OR. 4.16; 95% CI 4.05 to 4.28).

The most common comorbidities among the persons who were tested for SARS-CoV-2 at the PHCC testing sites were diabetes mellitus (11.2%) and hypertension (11.4%). The latter is a reflection of the noncommunicable diseases prevalence distribution among the PHCC target population. Based on the health needs assessment study that was conducted in 2019 among the PHCC registered population, the prevalence of diabetes mellitus ranged between 11.9% to 13.9%, and hypertension between 11.8% to 15.7%. ^18^

The highest positivity rate was observed among males coming from Asian countries at 20.3%. The latter might be attributed to their living conditions in more crowded areas and accommodations, and their social behavior of social mixing despite the national restrictions that limited individuals’ movement and gathering except in urgent situations.

This study applied a multivariable logistic regression model to understand the effect of effect sex, nationality group, age-groups, and symptoms presentation on capturing COVID-19 infection. Sex was found to have an effect with being a male was associated with a higher risk (OR 1.16; 95% CI 11.3 to 1.18). In a similar study conducted in Tianjin in China, males were found to be at higher risk of developing severe COVID-19 infection which highlighted the need for early COVID-19 testing and intervention among them. ^17^

The strengths of this study include all the tested cases at the community level where PHCC had testing sites, with all the tests conducted at a single lab. PHCC provided accessibility for COVID-19 testing at the community level by allocating 4 COVID-19 testing and holding centers and at least one testing room in the remaining 23 health centers. All the tests performed at the primary care were included, providing a robust estimation of the positivity rated among those tested. There are limitations to this study. The exact geographical location was not completely captured in the system and noncommunicable disease data was not completely recorded in the patient electronic medical record.

In conclusion, this study provides detailed information on the epidemic of COVID-19 in Qatar by analyzing the epidemiological characteristics of the tested cases at the primary health care level. The epidemic predominantly affected younger ages and males namely coming from Asia. At the primary health care level, the COVID-19 infection rate was higher among those who presented with clinical symptoms. The scale-up of the testing at the community level helped in detecting more cases and was reflected in a steady increase in the positivity rate to be flattened afterward.

## Data Availability

The data are avilabible upone request.

---

**Graph No. 1:**
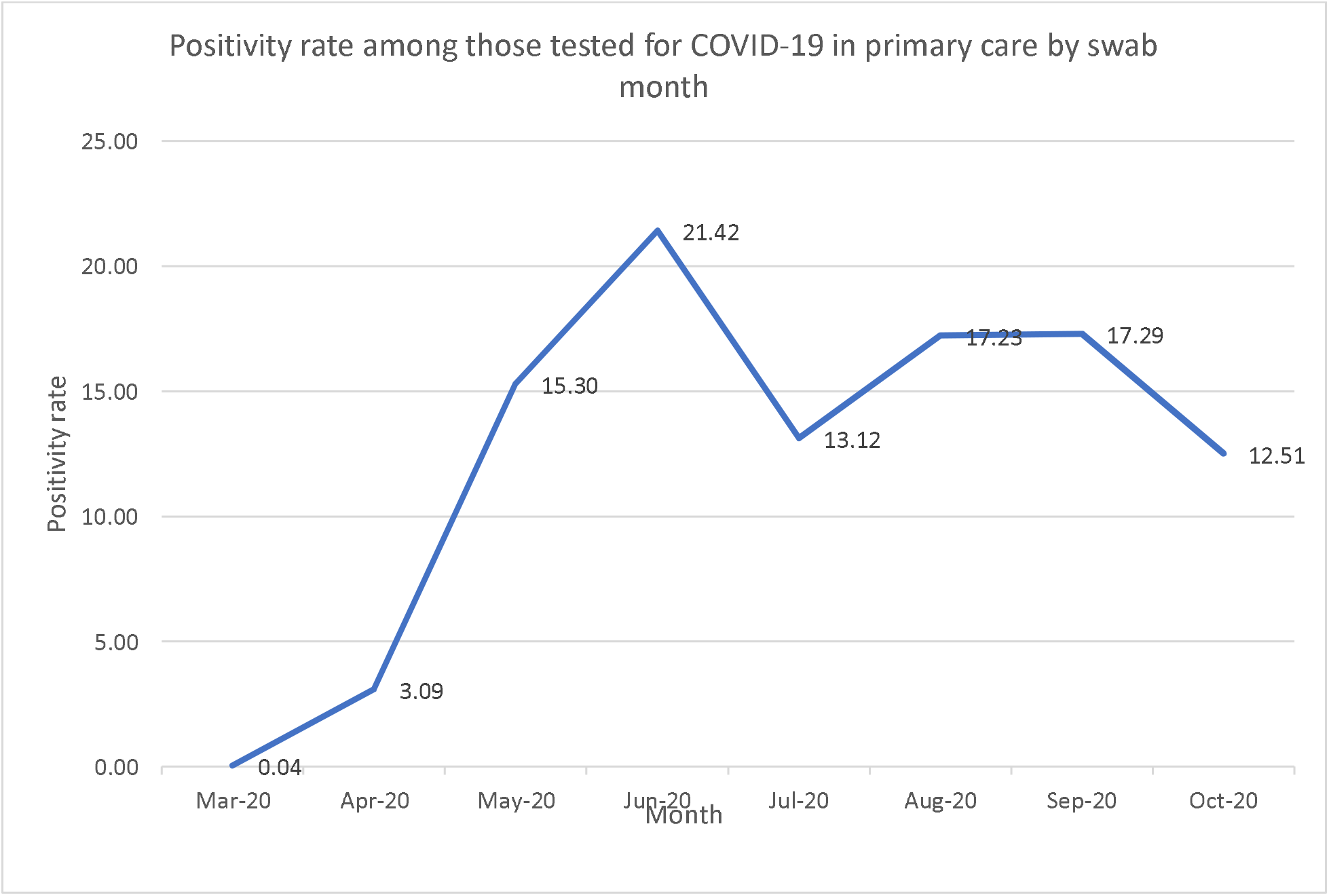
Positivity rate among those tested for COVID-19 in primary care by swab month.

## Notes

### Competing Interest Statement

The authors have declared no competing interest.

### Funding Statement

No funding support was requested for this study.

### Author Declarations

The Primary Health Care Corporation (PHCC) scientific research committee representing the PHCC IRB reviewed the research protocol and provide the required ethical approval.

## Reference

1. Zhu N, Zhang D, Wang W, et al. A novel coronavirus from patients with pneumonia in China, 2019. N Engl J Med 2020;382:727–33.

2. Chen N, Zhou M, Dong X, et al. Epidemiological and clinical characteristics of 99 cases of 2019 novel coronavirus pneumonia in Wuhan, China: a descriptive study. Lancet 2020;395:507–13.

3. Huage C, Wang Y, Li X. Clinical features of patients infected with 2019 novel coronavirus in Wuhan, China. Lancet 2020; 395: 497–506

4. Phan LT, Nguyen TV, Luong QC, et al. Importation and human-to-human transmission of a novel coronavirus in Vietnam. N Engl J Med 2020;382:872–4.

5. Young BE, Ong SWX, Kalimuddin S, et al. Epidemiologic features and clinical course of patients infected with SARS-CoV −2 in Singapore. JAMA 2020;323:1488.

6. Holshue ML, DeBolt C, Lindquist S, et al. First case of 2019 novel coronavirus in the United States. N Engl J Med 2020;382:929–36

7. WHO. WHO Coronavirus disease (COVID-19) dashboard. Geneva: World Health Organization, https://covid19.who.int/ (accessed June 9, 2020)

8. Oksanen A, Kaakinen M, Latikka R, et al. Regulation and trust: 3-month follow-up study on COVID-19 mortality in 25 European countries. JMIR Public Health Surveill 2020;6:e19218.

9. CIA. The World Factbook, 2020. https://www.cia.gov/the-worldfactbook/countries/qatar/#people-and-society (accessed Dec 23, 2020)

10. World population review, 2020. https://worldpopulationreview.com/countries/qatar-population (accessed Dec 28, 2020)

11. Wu C, Chen X, Cai Y, et al. Risk factors associated with acute respiratory distress syndrome and death in patients with coronavirus disease 2019 pneumonia in Wuhan, China. JAMA Intern Med 2020;180:934.

12. Zhou F, Yu T, Du R, et al. Clinical course and risk factors for mortality of adult inpatients with COVID-19 in Wuhan, China: a retrospective cohort study. Lancet 2020;395:1054–62.

13. Yang, H., Chen, D., Jiang, Q., & Yuan, Z. (2020). High intensities of population movement were associated with high incidence of COVID-19 during the pandemic. Epidemiology and Infection, 148, E177. doi:10.1017/S0950268820001703

14. Al Kuwari HM, et al. BMJ Open 2020;10:e040428. doi:10.1136/bmjopen-2020-040428

15. ThermoFisher. TaqPath COVID-19 Combo kit, 2020. Available: https://assets.thermofisher.com/TFS-Assets/LSG/manuals/MAN0019372_TaqPathCOVID-19_Kit_Australia_NZ_IFU.Pdf

16. Roche. Cobas® SARS-CoV-2 test, 2020. Available: https://www.diagnostics.roche.com/us/en/products/params/cobas-sars-cov-2-test.Html

17. Wang J, Li Z, Cheng X, et al. Epidemiological Characteristics, Transmission Chain, and Risk Factors of Sever Infection of COVID-19 in Tianjin, a Representative Municipality City of China. Front. Public Health 8:198. doi: 10.3389/fpubh.2020.00198

18. Primary Health Care Corporation 2020. Health Needs Assessment. Strategy Planning and Health Intelligence Directorate. Primary Health Care Corporation. Doha. 2020

